# Lymphocyte profiles after a first demyelinating event suggestive of multiple sclerosis reveal early monocyte and B cell alterations

**DOI:** 10.1101/2023.11.13.23298459

**Authors:** Cesar Alvarez-Gonzalez, Annika Wiedemann, Maria Schroeder-Castagno, Steven Schepanski, Susanna Asseyer, Claudia Chien, Joseph Kuchling, Judith Bellmann-Strobl, Klemens Ruprecht, Carmen Infante-Duarte, Thomas Dörner, Friedemann Paul

## Abstract

**Introduction:** Often, an isolated clinical event suggestive of CNS demyelination confers a risk of conversion to multiple sclerosis. In this study, we investigate lymphocyte profiles after a first clinical event suggestive of multiple sclerosis (MS), which could contribute to the current understanding of early inflammatory responses in this demyelinating disease.

**Methods:** Twenty treatment-naïve clinically isolated syndrome (CIS) patients and fifteen healthy participants were included in our assessment of lymphocyte profiles and B cell subsets using multicolour flow cytometry. Analysis was made at 3-6 months (Baseline), 12, and 24 months after a first clinical event. We also performed a sub-analysis of patients that received glatiramer acetate (GLAT) after their baseline visit up to 24 months after the first clinical event.

**Results:** Our analysis revealed monocyte and B cell differences between groups. Percentages of CD19^+^CD20^+^ B cells were lower in CIS patients compared to healthy individuals at baseline. Additionally, monocyte distribution among groups was different. A subgroup analysis of patients treated with GLAT (n= 10) showed an increased percentage of naïve (p<0.05) and memory pre-switched (p<0.01) B cells up to 24 months after their baseline visit compared to the untreated group (n= 10).

**Conclusion:** Our results showed early monocyte and B cell subsets alterations in pwCIS. Moreover, GLAT-treated patients showed an increased percentage of naïve and memory pre-switched B cells after 24 months of treatment. Further research is needed to elucidate the role of B cells and monocyte disturbances during inflammatory processes after a first clinically-MS suggestive event.

## 1. Introduction

Around 85% of patients with an acute neurological isolated clinical event (patients with clinically isolated syndrome or pwCIS) progress to multiple sclerosis (MS) (Lin et al., 2021; Pitt et al., 2022). Thus, the characterisation of lymphocyte subpopulations in the early stages of MS could – in conjunction with clinical and paraclinical data – contribute to our understanding of disease pathomechanisms and help develop biomarkers for treatment response and prognostication (Baker, Herrod, Alvarez-Gonzalez, Giovannoni, et al., 2017; Jager et al., 2008).

Evidence from animal models of MS have long suggested that MS is a CD4 Th1/Th17 - cell-mediated disease. Nonetheless, current evidence indicates that B cells, other lymphocytes subpopulations, and monocytes contribute likewise significantly to the pathogenesis of MS (Baker et al., 2019; Bittner et al., 2017; Gelfand et al., 2017; Lassmann, 2019; Roodselaar et al., 2021). For instance, lymphocytic investigation from anti-CD20 therapies and oral cladribine (CLAD) studies showed that the decrease in disease activity could be attributed to the depletion of B cells and that the effect on T cells might not be as relevant as previously claimed (Baker, Herrod, Alvarez-Gonzalez, Zalewski, et al., 2017; Cellerino et al., 2020; Sèze et al., 2023; Staun-Ram et al., 2018).

Here, we aim to investigate phenotypic patterns of lymphocyte subsets after a first clinical event suggestive of MS. We hypothesised that analysis of lymphocyte profiles in treatment-naïve pwCIS may contribute to the development of clinically applicable biomarkers for treatment response and disease course prognostication. In addition, in a small explorative sub-analysis in pwCIS, we compared lymphocyte subpopulations between two groups: pwCIS receiving glatiramer acetate (GLAT) and pwCIS who remained untreated.

## 2. Patients & Methods

### 2.1 Patients

Twenty treatment-naïve pwCIS (without MS disease-modifying treatment) from the Berlin CIS cohort (NCT01371071), an ongoing prospective observational study initiated at Charité – Universitätsmedizin Berlin in 2011, were selected to assess lymphocyte profiles and B cell subsets by using multicolour flow cytometry. The inclusion criteria, as described previously (Gieß et al., 2017), were: >18 years of age and a first clinical event suggestive of demyelination of the CNS, according to the McDonald 2010 criteria (Polman et al., 2011), within the 6 first months before cohort enrolment. Exclusion criteria were inability or unwillingness to provide informed consent, incomplete collection of blood samples for follow-up, and a history of any health condition or devices impeding immunological, clinical, and radiological examination. Moreover, sample selection was also limited to those individuals with sufficient availability of frozen peripheral blood mononuclear cells (PBMCs). All participants provided written informed consent and ethics approval for this study was given by the institutional review board of Charité – Universitätsmedizin Berlin (EA1/180/10).

During the visits, all patients underwent clinical and magnetic resonance imaging (MRI) assessments and blood extraction for PBMCs isolation. The first visit (baseline visit) was scheduled between 3-6 months after a first clinical event suggestive of demyelination of the CNS according to the McDonald 2010 criteria, and before administration of GLAT. The second visit was performed at around 12 months after the first clinical event, and the third visit at around 24 months after the first clinical event. Out of twenty pwCIS, ten received GLAT after baseline visit (due to signs of disease activity), whilst the other half remained untreated. PBMCs were isolated using Biocoll separating solution and stored in liquid nitrogen according to standard operating procedures (SOP) at NeuroCure Clinical Research Center, Charité – Universitätsmedizin Berlin. Frozen PBMCs from fifteen unmatched healthy donors (HDs) were used as controls. Disease activity was defined as the presence of new lesions in T2-weighted and/or contrast-enhancing MRI, and/or new clinical events suggestive of demyelination of the CNS, and/or confirmed deterioration on EDSS at 12 and/or 24 months of follow-up.

### 2.2 Immunophenotyping

Frozen PBMCs samples were thawed and subsequently stained and analysed by flow cytometry. Briefly, a set of markers to discriminate between lymphocytic profiles and B cell subsets was established: CD3, CD4, CD14, CD19, CD20, CD25, CD27, CD38, CD127, CD138, IgD, and HLA-DR. A live/dead fixable blue dead cell stain kit (Invitrogen) was used per the manufacturer’s instructions to identify dead cells. Cells were stained for 15 min at 4°C, washed and filtered before acquisition.

Stained samples were analysed on LSR Fortessa X-20 (LSRFortessa; BD Biosciences, USA). To ensure stable analysing conditions, flow cytometer setup and tracking beads from BD were used daily. Diva and FlowJo software (v10.4; BD, USA) were used to determine different subpopulations. Results are expressed as percentages. For details on antibodies and gating strategy (see **Supplementary Figure 1)**.

### 2.3 MRI acquisition and post-processing

All study subjects were scanned using 3 Tesla (Tim Trio; Siemens, Erlangen, Germany) MRI scanners at the Berlin Center for Advanced Neuroimaging. Pre- and post-contrast 3D T1- weighted magnetisation prepared rapid gradient-echo (MPRAGE) images (resolution 1 × 1 × 1 mm^3^; TR = 1,900 ms, TE = 3.03 ms, TI = 900 ms, flip angle 9°) and a 3D T2-weighted fluid -attenuated inversion recovery (FLAIR) images (resolution 1 × 1 × 1 mm^3^; TR = 6,000 ms, TE = 388 ms, TI = 2,100 ms, flip angle 120°) were acquired for each patient. MPRAGE and FLAIR were cropped, co-registered to MNI-152 space (https://fsl.fmrib.ox.ac.uk/fsl/fslwiki/Fslutils#Tools), N4-bias corrected (http://stnava.github.io/ANTs/) and linearly co-registered with each other using FSL FLIRT (Greve & Fischl, 2009). Longitudinal co-registration was performed by using the same FSL tools and each MPRAGE and FLAIR from follow-up scans per patient were co-registered to their baseline scan. T2-hyperintense and gadolinium contrast-enhancing lesions were segmented from FLAIR and post-contrast MPRAGE images manually using ITK-SNAP (Rasche et al., 2018) by 2 expert MRI technicians with more than 10 years of experience with multiple sclerosis lesion identification. Whole brain T2-hyperintense lesion count was extracted using FSL cluster.

### 2.4 Statistics

Statistical analysis was performed using JAMOVI for macOS (The jamovi project (2020). jamovi (v.2.5.1.0) [Computer Software]. Retrieved from https://www.jamovi.org) and Prism 8.4.2 (GraphPad, La Jolla, USA), and R version 4.3.2 (2023-10-31) Platform: aarch64- apple-darwin20 (64-bit) Running under: macOS Sonoma 14.4. Independent t-test and Mann-Whitney were used to compare percentages between two unpaired groups. A Shapiro-Wilk normality test was performed. A Fisher’s exact test was used to analyse any difference between categorical variables. Unless stated otherwise, data shows mean percentage and standard deviation (SD).

Furthermore, a sub-analysis of lymphocytes in GLAT-treated and untreated pwCIS was performed using Analysis of Covariance (ANCOVA). The ANCOVA model was applied to evaluate the difference in quantified cell populations between GLAT-treated and untreated pwCIS groups at 24 months post-baseline. To control for potential confounding effects from earlier time points, the model included both baseline and 12-month quantifications as covariates. This approach allowed us to isolate the treatment effect at 24 months while accounting for variations at baseline and changes over the first 12 months. The model was specified as follows: ‘outcome_24m ∼ baseline + outcome_12m + treatment’, where ‘outcome_24m’ represents the outcome measure at 24 months after baseline, ‘baselinè refers to the baseline measurement, ‘outcome_12m’ is the 12 months measurement, and ‘treatment’ indicates whether the participant received GLAT or not.

Prior to interpreting statistical differences between groups, the assumptions of ANCOVA, including normality of residuals using Shapiro-Wilk tests and homoscedasticity using Breusch-Pagan tests were formally tested. Data were defined as statistically significant when at least p < 0.05. Given small sample sizes, outlier detection was not performed, and thus not excluded from analysis.

## 3. Results

### 3.1 Demographics

Twenty pwCIS (male-to-female ratio 9:11) and fifteen HDs (male-to-female ratio 7:8) were included in this study (**Table 1**). No relevant difference between the two groups in sex (p=0.909) and age (p=0.548), was found. The baseline median EDSS for pwCIS was 2.00 (1.00 – 4.00) and 1.00 (0.00 – 2.50) for males and females, respectively. PBMCs from one CIS individual on baseline were not viable for flow cytometry analysis due to poor viability after thawing. Whilst in the GLAT group, disease activity was high at the baseline, no major differences in disease activity were found between groups at any time point afterwards (see Table 1 and **Supplementary Figure 2**).

**Table 1.** Demographic and clinical characteristics of pwCIS and healthy donors.

|  | HD | pwCIS<br>Baseline* | pwCIS<br>12 months* | pwCIS<br>24 months* |
| --- | --- | --- | --- | --- |
| Number, *n* | 15 | 20 | 20 | 20 |
| Untreated | - | 10 | 10 | 10 |
| GLAT | - | 10 | 10 | 10 |
| Age, years, median(*range*) | 38.2(21-64) | 32.5(24-47) | - | - |
| Untreated | - | 33.5(25-44) | - | - |
| GLAT | - | 31(24-47) | - | - |
| female/male, *n/n* | 8/7 | 11/9 | - | - |
| EDSS, median(*range*) | - |  |  |  |
| Untreated | - | 1.5(0-2.5) | 1.5(0-2.5) | 1.5(0-2) |
| GLAT | - | 1.5(0-4) | 1.5(0-4) | 2(0-4.5) |
| MRI T2-hyperintense lesion<br>count, median (*range*) |  |  |  |  |
| Untreated | - | 6(3-32) | 0(0-12) | 0(0-11) |
| GLAT | - | 25(6-52) | 2(0-11) | 1(0-6; n=9) |
| Relapses, median(*range*) |  |  |  |  |
| Untreated | - | - | 0(0-1) | 0(0-1) |
| GLAT | - | - | 0(0-1) | 0(0-1) |

HD healthy donor, pwCIS patients with clinical isolated syndrome (CIS), *a.b.* after baseline, *n* number, IQR interquartile range, “-“not applicable, EDSS Expanded Disability Status Scale, *range* (minimum - maximum values), GLAT glatiramer acetate, *Untreated* treatment naïve, \*At 3-6 (baseline), 12 and 24 months after baseline.

### 3.2 Differences between treatment-naïve pwCIS and HDs at baseline

First, we analysed the difference between lymphocytes and monocytes in pwCIS and HDs at baseline. Results are shown in **Figure 1** and **Table 2 (descriptive results in supplement table 2)**. We found notable differences that showed lower percentages of lymphocytes in pwCIS compared to HDs **(Figure 1A**; 84.0 vs 91.7; [**% mean** *p*□ 0.001]). In contrast, the frequency of monocytes was higher among pwCIS than HDs (**Figure 1A**; 16.0 vs 8.2,; [**% mean** *p* < 0.001]).

**Figure 1.**
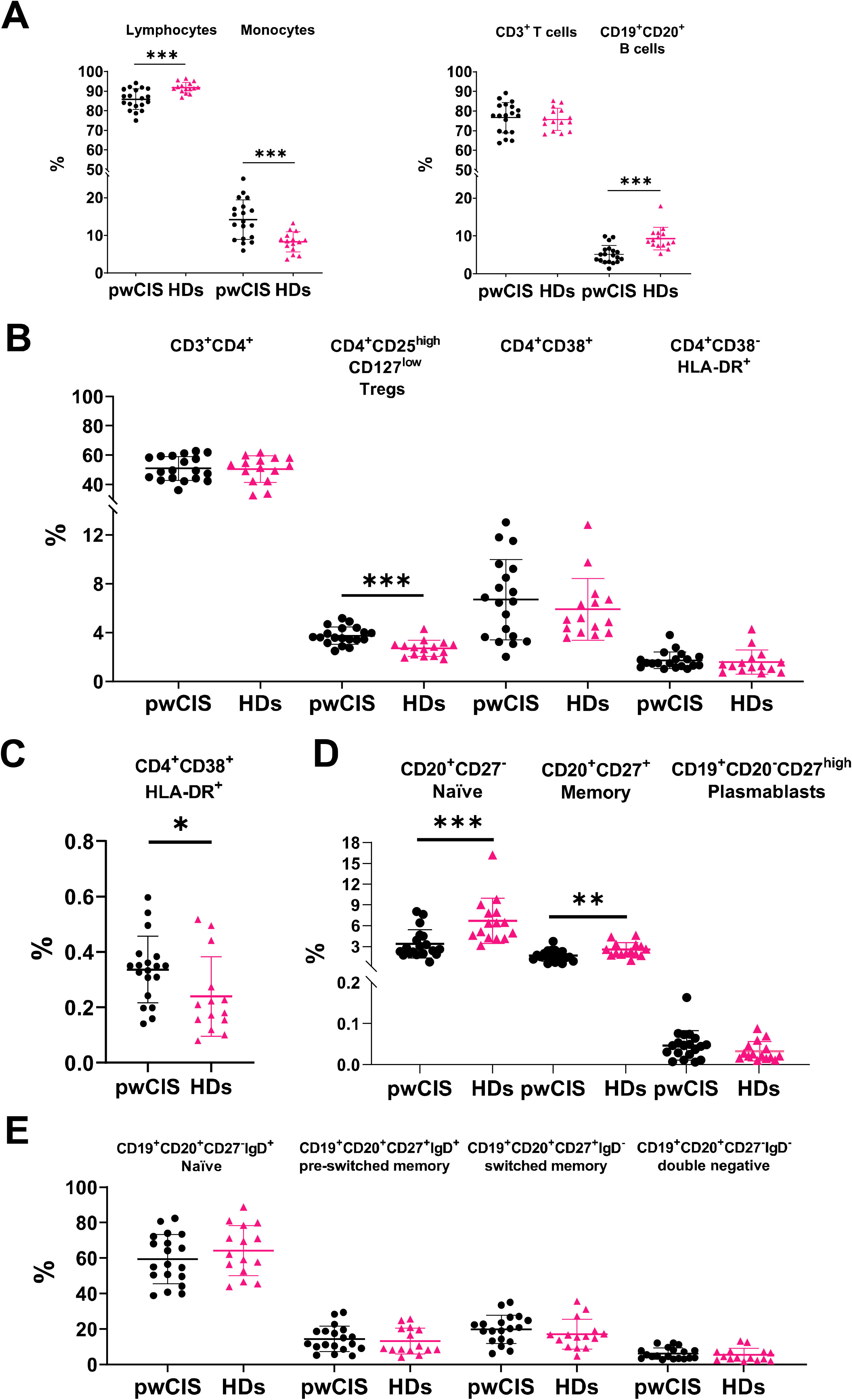
Differences between pwCIS and HDs at Baseline. A) Mean percentage of lymphocytes and monocytes between isolated PBMCs. Mean percentage of CD3^+^ T cells and CD19^+^CD20^+^ B cells in the lymphocyte population. B and C) Mean percentage of different T cell subpopulations in the lymphocyte population. D) Mean percentage of different B cell subpopulations in the lymphocyte population. E) Mean percentage of different B cell subpopulations in the CD20^+^ subpopulation. Data are presented as mean percentage ± SD. Mann-Whitney test was utilised to compare the percentages between two unpaired groups. \**p*□ 0.05, \*\**p*□ 0.01, \*\*\**p*□ 0.001.

**Table 2.** Differences between pwCIS and HDs at Baseline.

| Lymphocyte subsets | pwCIS<br>% (mean ±SD) *n*=19 | HDs<br>% (mean ±SD) *n*=15 | p-value |
| --- | --- | --- | --- |
| **CD3<sup>+</sup> (T cells)** | 76.6 (7.64) | 75.8 (5.62) | 0.56 |
| **CD4<sup>+</sup>** | 50.9 (8.21) | 50.3 (9.04) | 0.86 |
| CD4<sup>+</sup> CD38<sup>+</sup> | 6.68 (3.28) | 5.89 (2.52) | 0.63 |
| CD4<sup>+</sup> HLA-DR<sup>+</sup> | 1.72 (0.693) | 1.57 (0.996) | 0.25 |
| CD4<sup>+</sup>CD38<sup>+</sup>HLA-DR<sup>+</sup> | 0.33 (0.120) | 0.23 (0.140) | 0.015 |
| CD4<sup>+</sup><br>CD25<sup>high</sup>/CD127<sup>low</sup><br>(Tregs) | 3.75 (0.718) | 2.70 (0.652) | < 0.001 |
| **CD19<sup>+</sup>CD20<sup>+</sup> (B cells)** | 5.1 (2.37) | 9.32 (2.98) | < 0.001 |
| CD20<sup>+</sup>CD27<sup>-</sup><br>(Naïve) | 3.36 (2.04) | 6.68 (3.27) | < 0.001 |
| CD20<sup>+</sup>CD27<sup>+</sup><br>Memory B cells | 1.65 (0.792) | 2.55 (1.00) | 0.006 |
| CD19<sup>+</sup>CD20<sup>-</sup>CD27<sup>high</sup><br>Plasmablasts | 0.047 (0.0358) | 0.03 (0.0233) | 0.17 |
| B cell Subsets | pwCIS<br>% (mean ±SD) *n*=19 | HDs<br>% (mean ±SD) *n*=15 | p-value |
| CD20<sup>+</sup>CD27<sup>-</sup> IgD<sup>+</sup><br>Naïve | 59.45 (13.8) | 64.26 (14.1) | 0.354 |
| CD20<sup>+</sup>CD27<sup>+</sup>IgD<sup>+</sup><br>pre-switched memory | 14.43 (7.21) | 13.20 (7.29) | 0.471 |
| CD20<sup>+</sup>CD27<sup>+</sup>IgD<sup>-</sup><br>switched memory | 19.91 (7.92) | 17.13 (8.42) | 0.242 |
| CD20<sup>+</sup>CD27<sup>-</sup>IgD<sup>-</sup><br>double negative | 6.25 (3.19) | 5.40 (3.63) | 0.881 |

*Mann-Whitney test was utilised to compare the percentages between two unpaired groups.*

Lymphocyte subsets were categorised as shown in **Figure 1**. Briefly, T cells were defined by the expression of CD3, monocytes by the expression of CD14 and B cells characterised by the absence of CD3 and CD14 with the expression of CD19 and CD20. Afterwards, a set of markers to discriminate between specific T and B cell subsets was established (see gating strategy in **Supplement Figure 1**). Our results did not show any difference in the mean percentage of CD3^+^ (**Figure 1A**) or CD3^+^CD4^+^ T cells among groups (**Figure 1B**). However, the fraction of CD4^+^CD38^+^HLADR^+^ within the T cell population was slightly higher in pwCIS (**Figure 1C**; 0.36 vs 0.23; [**% mean** *p*=0.015]). Moreover, the percentage of CD4^+^CD25^high^/CD127^low^ cells (defining Tregs) in pwCIS was somewhat higher than in HDs **(Figure 1B**; 3.9 vs 2.6; [**% mean** p□ 0.001]).

Interestingly, the percentage of CD19^+^CD20^+^ B cells among lymphocytes was notably lower in pwCIS compared to HDs (**Figure 1A**; 5.7 vs 9.3; [**% mean** p□ 0.001]). Percentages of both, CD20^+^CD27^-^ naïve and CD20^+^CD27^+^ memory B cells, were higher in HDs (naïve= 4.0 and memory= 2.5; [**% mean**]) than in pwCIS (naïve= 6.6 and memory= 1.5; [**% mean**]) (**Figure 1D**; *p*□ 0.001 and *p*□ 0.01, respectively); however, there was no substantial difference between the percentages of CD19^+^CD20^-^CD27^high^ cells (Plasmablasts; **Figure 1D**).

When we compared the proportions of B cell subsets defined by CD27 and IgD markers (CD27^-^IgD^+^ naïve, CD27^+^IgD^+^ pre-switched memory, CD27^+^IgD^-^ switched memory, and CD27^-^IgD^-^ double negative) between pwCIS and HDs, we did not find any substantial difference among these cohorts (**Figure 1E**).

### 3.3 Sub-analysis of lymphocytes in treated-GLAT and untreated pwCIS

Using ANCOVA analyses, we compared lymphocyte subsets and monocytes in two subgroups of patients: those who were untreated (n=10) and those who received glatiramer acetate (GLAT; n=10). While controlling for baseline (3-6 months) and 12-month quantifications as covariates, the outcomes analysed for the two subgroups of patients were at 24 months after the first clinical event suggestive of demyelination of CNS. Results from ANCOVA analyses are depicted in **Table 3 (descriptive results in Supplement Table 3 and Supplement Figures 4 and 5)**.

**Table 3.**
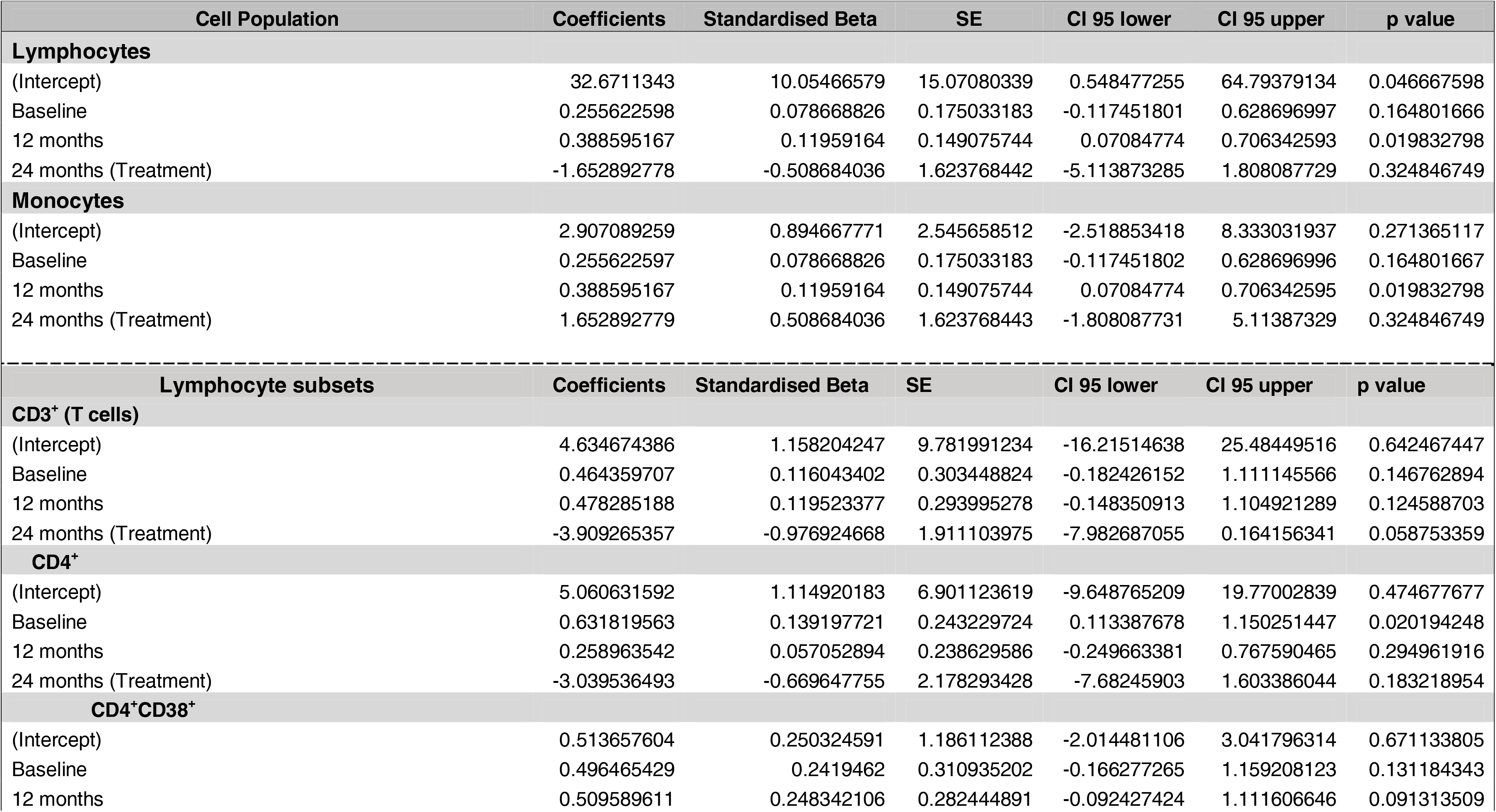
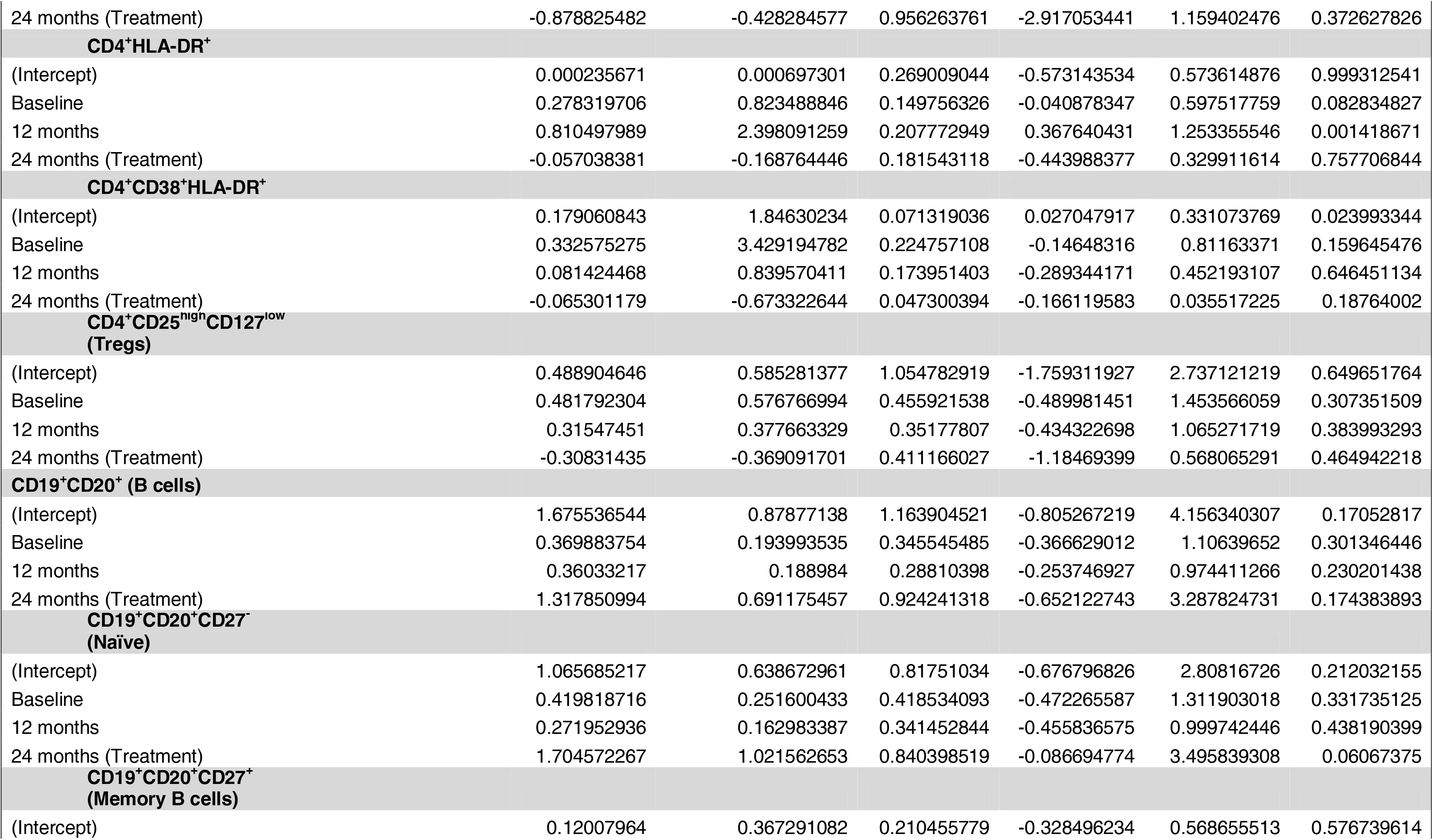
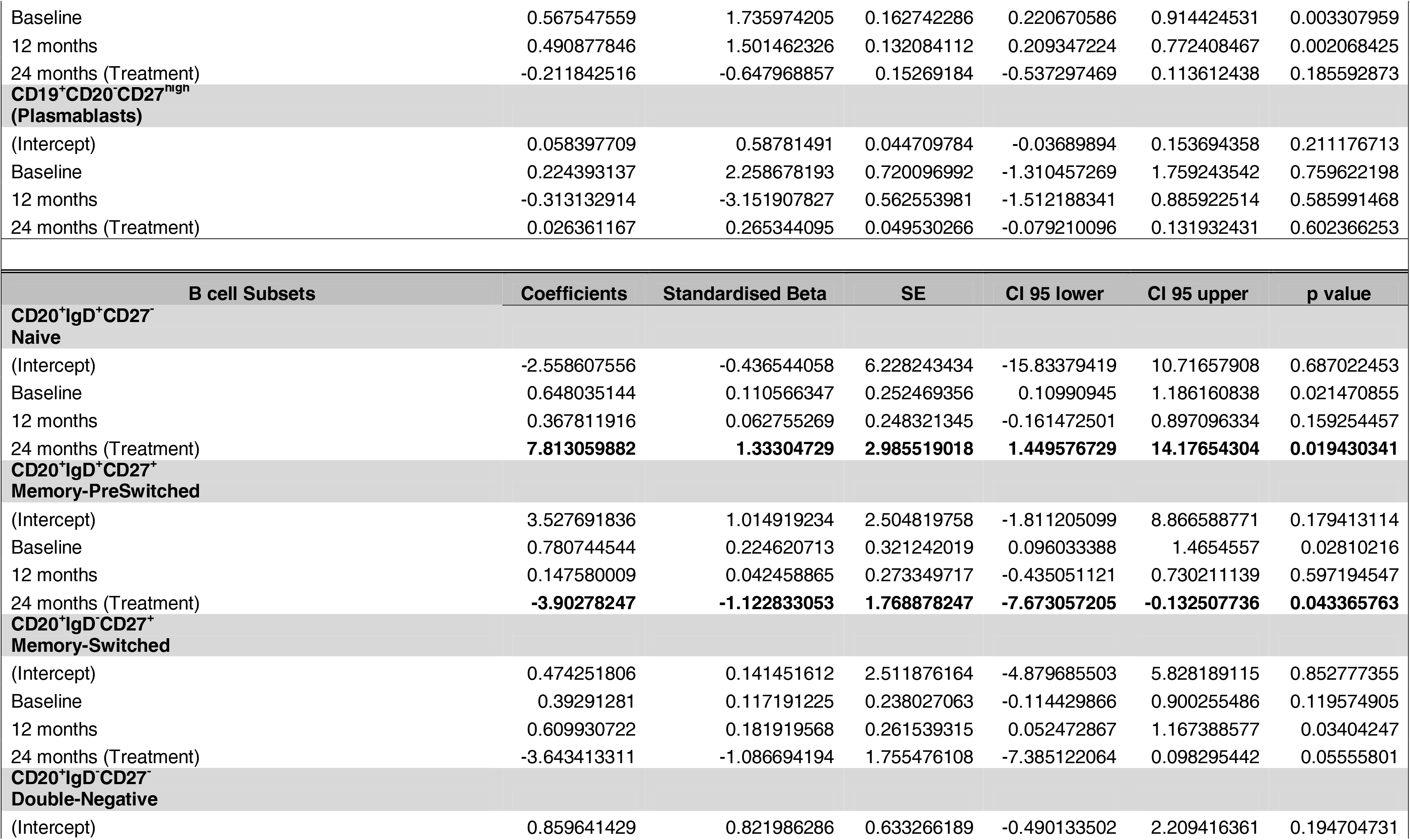
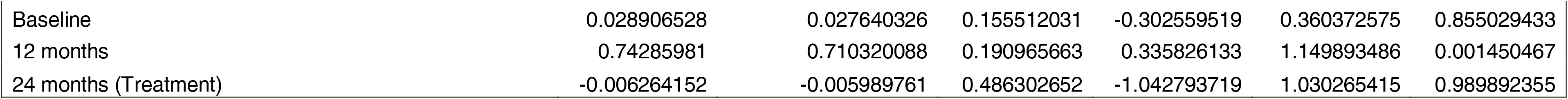
ANCOVA model coefficients for predicting 24-months cell populations.

First, we analysed the difference between lymphocytes and monocytes in pwCIS at 24 months. We, nonetheless, did not find any important changes among groups.

We categorised lymphocyte subsets as shown in **Table 3** (see also gating strategy in **Supplement Figure 1**). Our analysis showed no difference in CD3^+^ T cells and their subpopulations at 24 months between untreated and GLAT-treated pwCIS. Our results also showed that the percentage of CD19^+^CD20^+^ B cells were present at similar frequencies at baseline for both patient groups. However, while B cells increased at 24 months (GLAT=7.85 and Untreated= 5.47; [**% mean**]) in the GLAT-treated group, no significant difference was found (p=0.17). This non-statistically significant difference in the GLAT group appears to be secondary to an increased percentage of CD20^+^CD27^-^ naïve B cells (*p=* 0.06). The percentage of CD20^+^CD27^+^ memory B cells was slightly reduced in the GLAT group at 24 months, whilst the untreated group had a little increase, but no significant difference was observed (p=0.18). No significant differences were observed in plasmablast percentages at 24 months.

Finally, we compared the proportions of B cell subsets (CD27^-^IgD^+^ naïve, CD27^+^IgD^+^ pre-switched memory, CD27^+^IgD^-^ switched memory, and CD27^-^IgD^-^ double negative) between untreated and GLAT-treated patients. Interestingly, we confirmed the tendency of the percentage of CD20^+^CD27^-^IgD^+^ naïve B cells to increase within the group under GLAT after 24 months (*p*= 0.01). Our results also showed that the percentage of CD20^+^CD27^+^IgD^+^ pre-switched memory B cells in the GLAT group was lower at 24 months (*p*= 0.04); in stark contrast to the GLAT group, pre-switched memory B cells tended to increase in the untreated group compared to baseline. In the GLAT group, the percentage of CD20^+^CD27^+^IgD^-^ switched memory B cells decreased at 24 months, but it was not statistically significant (p=0.056). Lastly, our analysis did not show any substantial change in the percentage of CD20^+^CD27^-^IgD^-^ double negative B cell subpopulation.

## 4. Discussion

Our observational explorative study aimed to characterise lymphocyte subsets after a first demyelinating event suggestive of MS. Our results showed early monocyte and B cell alterations in pwCIS using a broad immunophenotyping platform.

### 4.1 Differences between pwCIS and HDs

Results showed that the distribution of peripheral lymphocytes and monocytes differs between HDs and pwCIS. The percentage of lymphocytes in pwCIS is lower whilst the percentage of monocytes is higher compared to healthy individuals. Previous studies have highlighted the important role of monocytes in MS pathogenesis; they can contribute to the breakdown of the blood-brain barrier (Waschbisch et al., 2016). Furthermore, the T cell/monocyte ratio analysis has identified pwCIS at risk of rapid disease progression (Nemecek et al., 2016). In line with previous studies, our results encourage further characterisation of monocyte subsets (in connection with dynamic changes among B cell subsets) in pwCIS in future clinical research projects with large samples.

Within the T cell compartment, the distribution of CD4^+^ cells did not show differences between HDs and pwCIS. Specifically, CD4^+^ subpopulations expressing activation markers, such as CD38 and HLA-DR, did not differ from the aforementioned groups. Interestingly, the proportion of T regulatory cells (Tregs) was higher in pwCIS compared to HDs at baseline. However, a steady, higher proportion of peripheral Tregs did not seem to show any further benefit in preventing disease activity during the 24 months of follow-up; to confirm this tendency, it is necessary to conduct additional studies with a larger sample size.

Moreover, increasing evidence shows that several cell subsets contribute to the inflammatory cascade inside the CNS (Büdingen et al., 2012; Lisak et al., 2017; Stern et al., 2014). Our results showed that the percentage of CD19^+^CD20^+^ B cells was lower in pwCIS compared to HDs. This is an interesting finding, considering that within the T cell compartment there was no or very little difference between groups. Our results are in line with Kreuzfelder *et al*. which showed that the percentage of peripheral CD19^+^ cells in pwMS is lower than in HDs (Kreuzfelder et al., 2004). Whether this relatively low percentage of peripheral B cells is due to a higher monocyte proportion in pwCIS or to an active exchange between the periphery and CNS, including an increased influx of B cells into the inflamed CNS tissue, needs further investigation (Stern et al., 2014).

Intriguingly, a separate analysis of B cell subsets (with the CD19^+^CD20^-^ plasmablast population excluded,) showed that the percentage of peripheral switched memory, pre-switched memory, and double negative B cell subpopulations are slightly higher in pwCIS, but the difference is not statistically significant; further studies with a larger sample size are needed to confirm this tendency.

### 4.2 Sub-analysis in pwCIS receiving GLAT

We also performed an ANCOVA analysis, as previously described, on two groups: untreated pwCIS and GLAT-treated pwCIS. Taken as a whole, lymphocytes and monocytes, we did not observe any significant change between those untreated compared to GLAT-treated individuals. We, nonetheless, observed a statistically non-significant increase in the percentage of B cells (among lymphocyte subsets) in those patients being treated with GLAT. Our results showed no significant changes within the plasmablast population (CD19^+^CD20^-^CD27^high^) subset.

Among B cell subsets, our analysis showed a higher percentage of naïve (*p*□ 0.01) and lower pre-switched memory (*p*□ 0.04) B cells after 24 months in the GLAT group, compared to the untreated group. Furthermore, whilst the percentage of switched memory B cells seemed to be reduced after 24 months in people receiving GLAT treatment, our analysis did not show a substantial change among groups at different time points (p=0.055); it would be worth exploring this outcome in further studies with a larger sample size, however.

Due to this study’s explorative nature and aim, it is difficult to make assumptions about the therapeutic effect of GLAT in pwCIS, and therefore, our results are just observational; interindividual differences can also drive these changes. Interestingly, it has been observed that GLAT could affect various aspects of dysregulated B cells subsets in pwMS (Ireland et al., 2014; Kemmerer et al., 2020). Moreover, data from Häuser et al. showed that GLAT modulates B cells by downregulating the activation markers CD69, CD95, and CD25, decreasing TNF-α production, and increasing IL-10 secretion. GLAT also expands CD4^+^CD25^+^FoxP3^+^ Treg cells (Häusler et al., 2020).

### 4.3 B cells and immune profiles in pwCIS and MS

Whilst B cells could overreact, generating a hostile and extreme autoimmune reaction, evidence has also shown that some subsets could regulate autoreactive T cell responses (Rieger & Bar-Or, 2008; Staun-Ram & Miller, 2017). For instance, it is believed that alemtuzumab depletes CD19^+^CD27^+^ B memory cells and allows a rapid repopulation of immature B cells (Baker, Herrod, Alvarez-Gonzalez, Giovannoni, et al., 2017). Furthermore, a new study by Ruck *et al*., using an in-depth multidimensional immune phenotyping analysis, showed that the presence of hyperexpanded T cell clones might predict the development of secondary autoimmunity in some patients under alemtuzumab treatment. The effect of these T cell clones in MS pathophysiology is still unclear (Ruck et al., 2022). The depletion of peripheral B cells by anti-CD20 therapies shows that B cells play a key role in MS pathogenesis (Comi et al., 2021). However, the depletion of B cell subsets might not be enough to maintain an effective therapeutic outcome. Depletion of other reactive T cell subsets to obtain a more effective response might be needed. (Disanto et al., 2012; Graf et al., 2020) Moreover, some studies have found that the production of interleukin-10 (IL-10) by B cell subsets is a key factor for regulatory responses (Fillatreau et al., 2002, 2008; Rieger & Bar-Or, 2008). It has been proposed that IL-10 produced by naïve and memory B cells has a direct effect on activated Th1/Th17 and T regulatory cells to regulate immune responses in both health and pathological conditions (Fillatreau et al., 2008). Further research is necessary to determine if lymphocyte immunophenotyping profiles in pwCIS can predict disease progression and/or guide personalised treatment decisions.

It is of note that we were unable to observe plasma cells (expression of CD138) in all measured samples. It seems that temporary circulating plasma cells are less likely to survive the freezing/thawing procedures applied to PBMCs. Moreover, in our study, we did not perform absolute counts due to the use of frozen samples. We propose that future lymphocyte immunophenotyping shall be performed in fresh blood and include absolute counting. This would not have been possible with the current methodology of this cohort. Moreover, it is important to mention that the decision to include only pwCIS using GLAT was influenced by factors beyond our control of this cohort; at the time of enrolment, GLAT was one of the most widely prescribed drugs to treat CIS/early MS.

Nevertheless, despite the limitations of this exploratory observational retrospective study and small sample size, we consider our results could encourage further research by using methodological assets of phenotypic and molecular studies to identify cellular biomarkers in these MS entities. In this study, we show that lymphocyte characterisation may be useful in characterising differences in peripheral lymphocyte subsets in pwCIS after a first demyelinating event.

## 5. Conclusion

In conclusion, our results showed early monocyte and B cell subsets alterations in pwCIS. Therefore, in future extensive clinical research, lymphocyte alterations found in our study could be considered to examine potential associations with ongoing MS activity after a first clinical manifestation. In addition, further inclusion of a wider immunophenotyping platform in larger cohorts could help to identify predictive immunological patterns.

## Data Availability

The datasets generated and/or analysed during the current study are not publicly available due to local regulations concerning the protection of patient data, but upon reasonable request, approval for distribution of data will be obtained from the institutional review board of Charite-Universitatsmedizin Berlin and anonymised data will be made available by the corresponding authors.

## Acknowledgements

We thank patients and healthy donors who participated in this study. We thank Rene M. Gieß, Bibiane Seeger-Schwinge, Gritt Stoeffels, Claudia Messelhauser and Katharina Stoesslein (NeuroCure Clinical Research Center, ECRC, Charité - Universitätsmedizin Berlin, Germany) for their technical assistance.

## Availability of data and materials

The datasets generated and/or analysed during the current study are not publicly available due to local regulations concerning the protection of patient data, but upon reasonable request, approval for distribution of data will be obtained from the institutional review board of Charité - Universitätsmedizin Berlin and anonymised data will be made available by the corresponding authors.

## Authors’ contributions

CAG, AW, MSC, SS, CID, TD, FP: Conceived, designed the experiments and interpreted the data; CAG, AW: Performed the experiments; CAG, AW, MSC: Analysed the data; CAG, SS, CID, SA, JK, JB-S, CC, KR: Helped with patients’ recruitment and clinical, MRI and/or laboratory data acquisition; CAG wrote the manuscript; CAG, CC, AW, MSC, SS, CID, TD, FP: Critically edited the manuscript.

