## Supplementary Data for "Lymphocyte profiles after a first demyelinating event suggestive of multiple sclerosis reveal early monocyte and B cell alterations"

### Supplement Table 1

#### Staining at Fortessa→B cells in MS

| target of antibody | clone | fluorochrome | manufacturer |
|---|---|---|---|
| **CD3** | UCHT1 | PE-Cy7 | BD Biosciences, Heidelberg, Germany |
| **CD4** | RPA-T4 | APC-Cy7 | BD Biosciences, Heidelberg, Germany |
| **CD14** | M5E2 | BV421 | Biolegend, San Diego, USA |
| **CD19** | SJ25C1 | BV711 | BD Biosciences, Heidelberg, Germany |
| **CD20** | 2H7 | BV510 | Biolegend, San Diego, USA |
| **CD25** | M-A251 | PE-CF594 | BD Biosciences, Heidelberg, Germany |
| **CD27** | M-T271 | APC-R700 | BD Biosciences, Heidelberg, Germany |
| **CD38** | HIT2 | PerCpCy5.5 | BD Biosciences, Heidelberg, Germany |
| **CD127** | HIL-7R-M21 | BV786 | BD Biosciences, Heidelberg, Germany |
| **CD138** | MI15 | BUV737 | BD Biosciences, Heidelberg, Germany |
| **IgD** | IA6-2 | PE | BD Biosciences, Heidelberg, Germany |
| **HLA-DR** | L243 | FITC | Biolegend, San Diego, USA |

Alvarez C., Wiedemann A., Schroeder-Castagno M., *et al.* **Lymphocyte profiles after a first demyelinating event suggestive of multiple sclerosis reveal early monocyte and B cell alterations**

### Supplement Figure 1: Gating Strategy

#### A) Lymphocytes and Monocytes

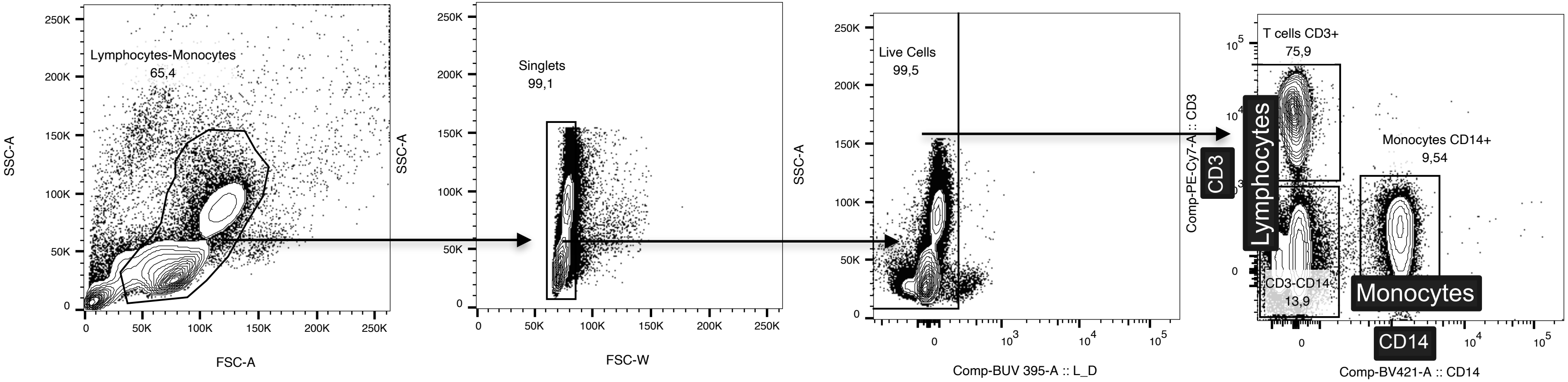


Gating for lymphocytes and monocytes in FSC-A and SSC-A

Gating for singlets

Gating for live events

Gating for CD3 and CD14 positive events

Alvarez C., Wiedemann A., Schroeder-Castagno M., *et al.* **Lymphocyte profiles after a first demyelinating event suggestive of multiple sclerosis reveal early monocyte and B cell alterations**

### B) CD3+ T cells and subsets

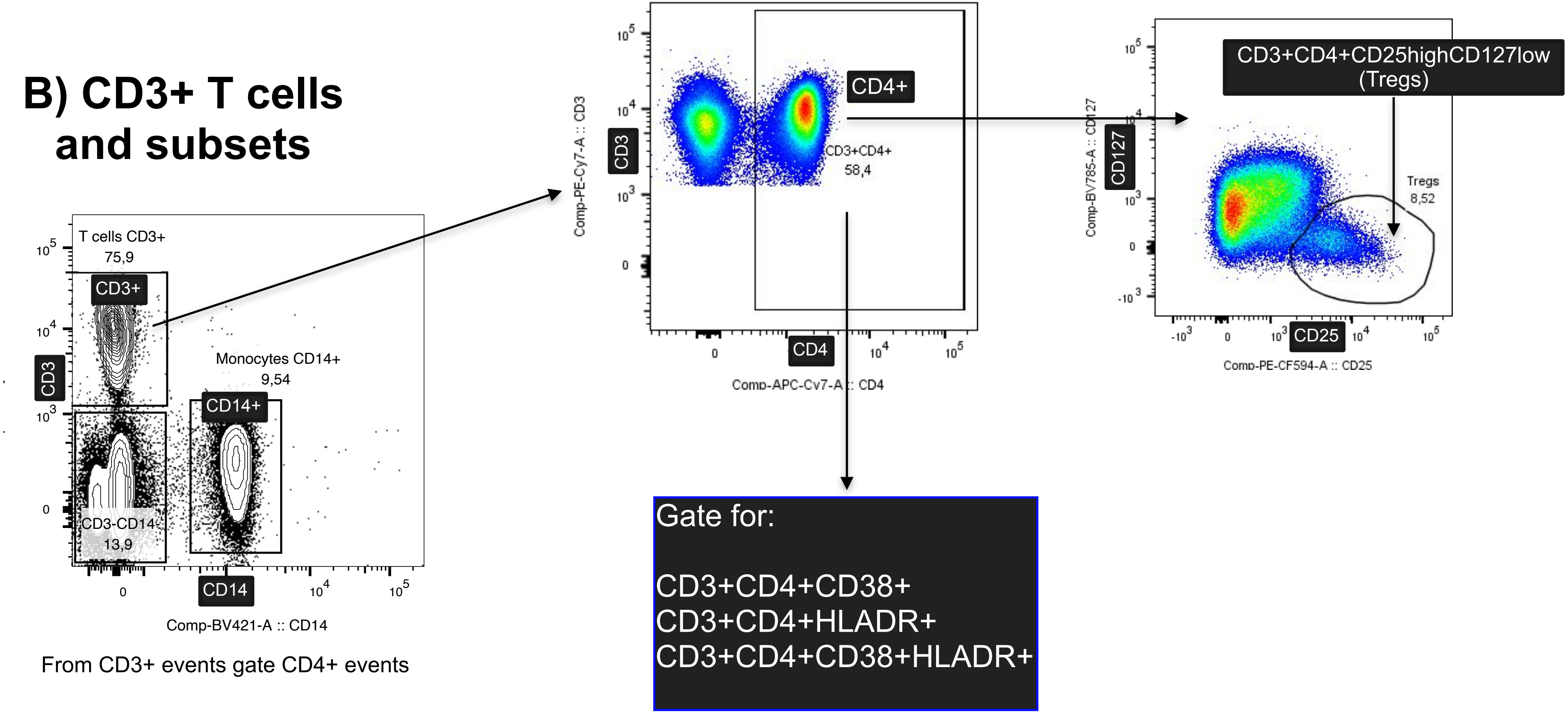


From CD3+ events gate CD4+ events

Alvarez C., Wiedemann A., Schroeder-Castagno M., *et al.* **Lymphocyte profiles after a first demyelinating event suggestive of multiple sclerosis reveal early monocyte and B cell alterations**

### C) CD19+CD20+ B cells and subsets

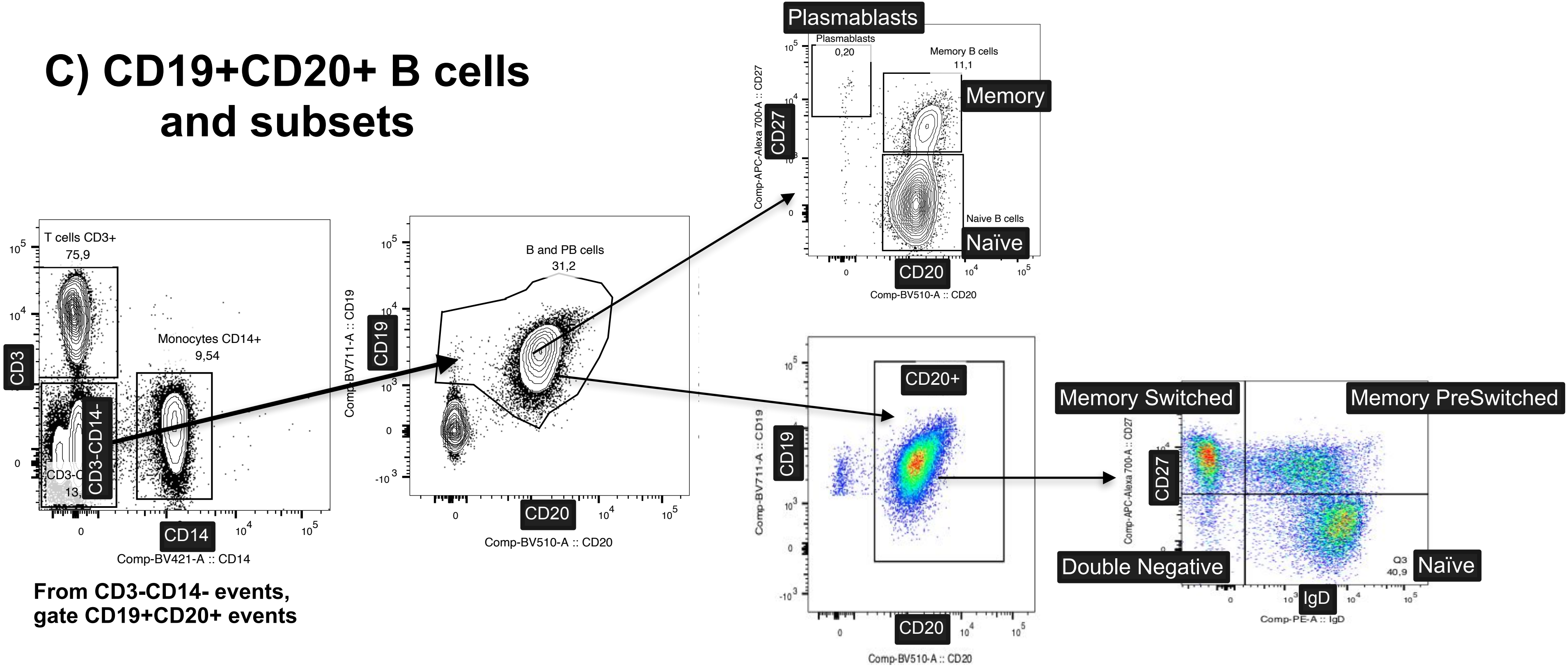


**From CD3-CD14- events, gate CD19+CD20+ events**

Alvarez C., Wiedemann A., Schroeder-Castagno M., *et al.* **Lymphocyte profiles after a first demyelinating event suggestive of multiple sclerosis reveal early monocyte and B cell alterations**

### Supplement Figure 2: Disease Activity

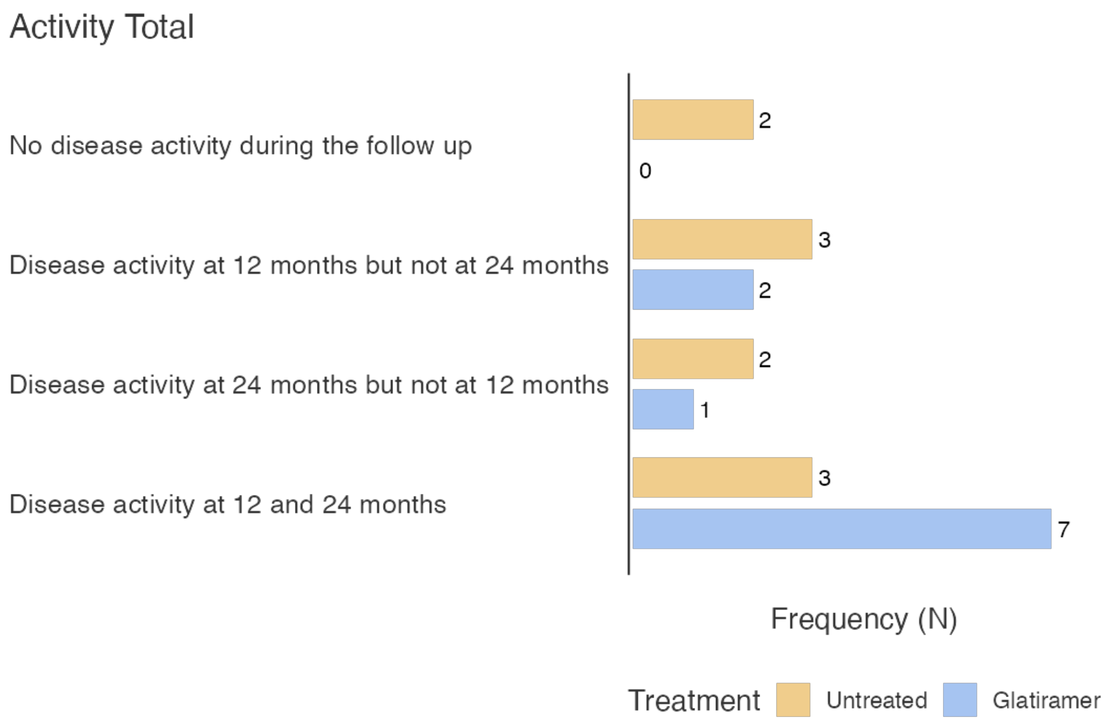


Alvarez C., Wiedemann A., Schroeder-Castagno M., *et al.* **Lymphocyte profiles after a first demyelinating event suggestive of multiple sclerosis reveal early monocyte and B cell alterations**

**Supplement Table 2.** Descriptive results of pwCIS and HDs at Baseline

| Cell Population (%) | Groups | N | Mean | SE | Median | SD |
|---|---|---|---|---|---|---|
| % of Lymphocytes | pwCIS | 19 | 85.7934 | 1.20292 | 86.3150 | 5.2434 |
| | HDs | 15 | 91.7422 | 0.69271 | 91.5025 | 2.6829 |
| % of Monocytes | pwCIS | 19 | 14.2066 | 1.20292 | 13.6850 | 5.2434 |
| | HDs | 15 | 8.2578 | 0.69271 | 8.4975 | 2.6829 |
| % of CD3+ (T cells) | pwCIS | 19 | 76.6404 | 1.75204 | 77.6206 | 7.6370 |
| | HDs | 15 | 75.7543 | 1.45014 | 74.4667 | 5.6164 |
| % of CD4+ | pwCIS | 19 | 50.9302 | 1.88247 | 49.3898 | 8.2055 |
| | HDs | 15 | 50.3401 | 2.33512 | 52.5042 | 9.0439 |
| % of CD4+CD38+ | pwCIS | 19 | 6.6866 | 0.75167 | 6.5443 | 3.2765 |
| | HDs | 15 | 5.8940 | 0.65072 | 5.0397 | 2.5202 |
| % of CD4+ CD38- HLA-DR+ | pwCIS | 19 | 1.7218 | 0.15900 | 1.4946 | 0.6931 |
| | HDs | 15 | 1.5770 | 0.25707 | 1.2590 | 0.9956 |
| % of CD4+ CD38+ HLA-DR+ | pwCIS | 19 | 0.3359 | 0.02760 | 0.3484 | 0.1203 |
| | HDs | 15 | 0.2328 | 0.03627 | 0.1794 | 0.1405 |
| % of CD4+ CD25high CD127low (Tregs) | pwCIS | 19 | 3.7478 | 0.16471 | 3.5994 | 0.7180 |
| | HDs | 15 | 2.6959 | 0.16826 | 2.7913 | 0.6517 |
| % of CD19+CD20+ (B cells) | pwCIS | 19 | 5.1218 | 0.54334 | 4.8820 | 2.3684 |
| | HDs | 15 | 9.3180 | 0.76821 | 8.4420 | 2.9752 |
| % of CD20+ CD27- Naïve | pwCIS | 19 | 3.3571 | 0.46839 | 2.5883 | 2.0417 |
| | HDs | 15 | 6.6776 | 0.84367 | 6.3229 | 3.2675 |
| % of CD20+ CD27+ Memory | pwCIS | 19 | 1.6519 | 0.18176 | 1.4935 | 0.7923 |
| | HDs | 15 | 2.5450 | 0.25944 | 2.2394 | 1.0048 |
| % of CD19+ CD20- CD27high Plasmablasts | pwCIS | 19 | 0.0472 | 0.00822 | 0.0465 | 0.0358 |
| | HDs | 15 | 0.0326 | 0.00601 | 0.0225 | 0.0233 |
| % of CD19+CD20+CD27-IgD+ Naïve | pwCIS | 19 | 59.4501 | 3.17569 | 56.5424 | 13.8425 |
| | HDs | 15 | 64.2632 | 3.65156 | 62.0955 | 14.1424 |
| % of CD19+CD20+CD27+IgD+ pre-switched memory | pwCIS | 19 | 14.4304 | 1.65485 | 11.6766 | 7.2133 |
| | HDs | 15 | 13.2042 | 1.88216 | 8.9897 | 7.2896 |
| % of CD19+CD20+CD27+IgD- switched memory | pwCIS | 19 | 19.9121 | 1.81677 | 20.2467 | 7.9191 |
| | HDs | 15 | 17.1319 | 2.17346 | 15.9067 | 8.4178 |
| % of CD19+CD20+CD27-IgD- double negative | pwCIS | 19 | 6.2499 | 0.73292 | 4.8285 | 3.1947 |
| | HDs | 15 | 5.4006 | 0.93733 | 3.9471 | 3.6303 |

#### Supplement Figure 3

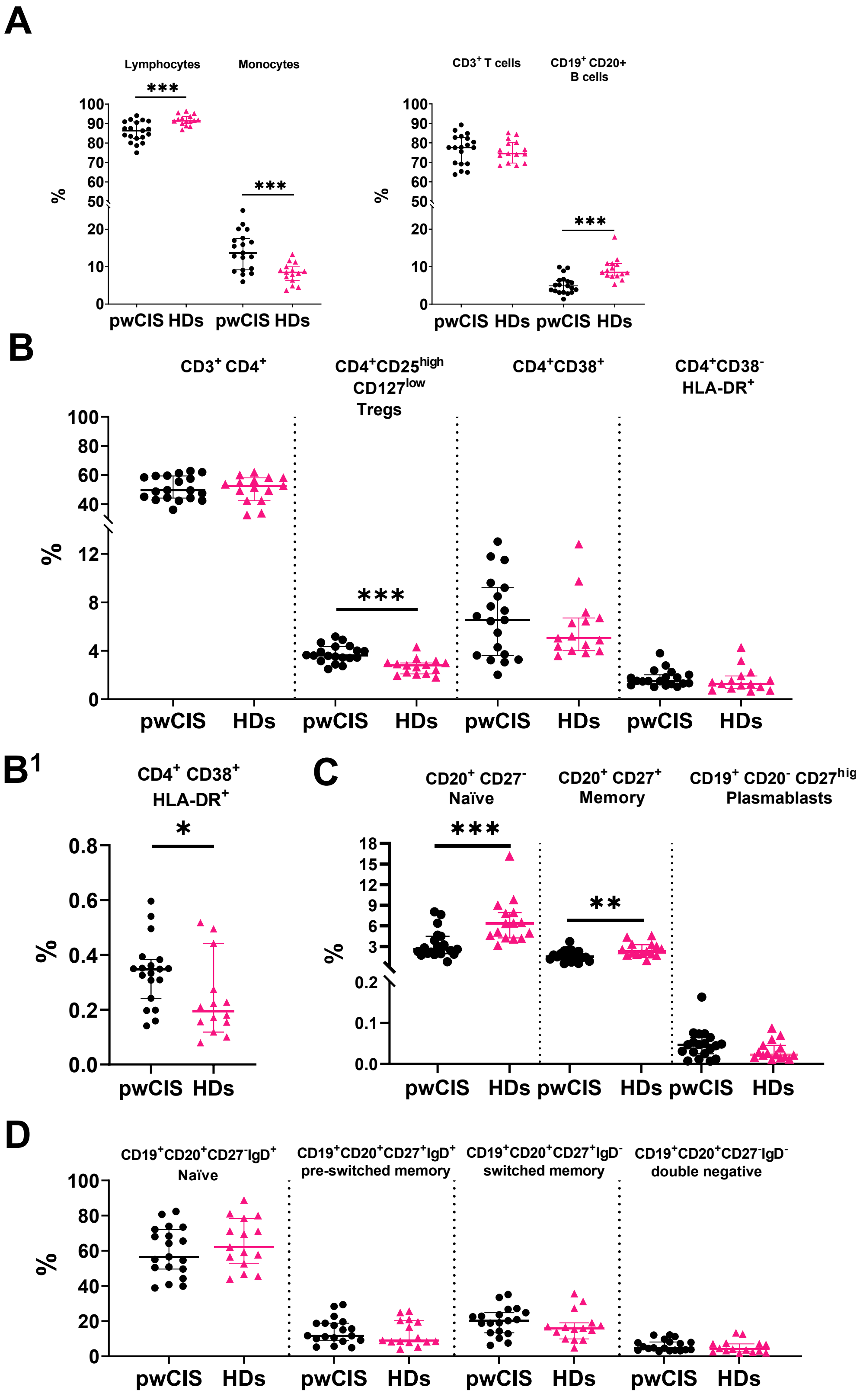


**Supplement Figure 3. Differences between pwCIS and HDs at Baseline.** A) Median percentage of lymphocytes and monocytes between isolated PBMCs. Median percentage of $CD3^{+}$ T cells and

CD19$^+$CD20$^+$ B cells in the lymphocyte population. B and C) Median percentage of different T cell subpopulations in the lymphocyte population. D) Median percentage of different B cell subpopulations in the lymphocyte population. E) Median percentage of different B cell subpopulations in the CD20$^+$ subpopulation. Data are presented as median percentage 95% CI. *$p < 0.05$, **$p < 0.01$, ***$p < 0.001$. Mann-Whitney test was utilised to compare the percentages between two unpaired groups.

**Supplement Table 3.** Descriptive results of sub-analysis of lymphocytes in treated-GLAT and untreated pwCIS

| Cell Population (%) | Treatment | N | Mean | SE | Median | SD |
|---|---|---|---|---|---|---|
| Lymphocytes (Baseline) | Untreated | 9 | 87.7862 | 1.61045 | 87.5507 | 4.8314 |
| | Glatiramer | 10 | 84.0000 | 1.63379 | 83.9972 | 5.1665 |
| CD3+ (Baseline) | Untreated | 9 | 77.6213 | 3.11314 | 78.9008 | 9.3394 |
| | Glatiramer | 10 | 75.7575 | 1.93094 | 77.5435 | 6.1062 |
| CD4+ (Baseline) | Untreated | 9 | 48.8100 | 2.81079 | 48.5799 | 8.4324 |
| | Glatiramer | 10 | 52.8384 | 2.50829 | 53.4231 | 7.9319 |
| CD4+CD38+ (Baseline) | Untreated | 9 | 6.3664 | 1.04954 | 5.4899 | 3.1486 |
| | Glatiramer | 10 | 6.9747 | 1.11639 | 6.6955 | 3.5303 |
| CD4+HLADR+ (Baseline) | Untreated | 9 | 1.4417 | 0.14277 | 1.3196 | 0.4283 |
| | Glatiramer | 10 | 1.9739 | 0.25466 | 1.7829 | 0.8053 |
| CD4+CD38+HLARDR+ (Baseline) | Untreated | 9 | 0.3014 | 0.03958 | 0.3092 | 0.1187 |
| | Glatiramer | 10 | 0.3670 | 0.03760 | 0.3505 | 0.1189 |
| CD4+CD25highCD127low: Tregs (Baseline) | Untreated | 9 | 3.5116 | 0.20725 | 3.5747 | 0.6218 |
| | Glatiramer | 10 | 3.9603 | 0.24111 | 3.6189 | 0.7625 |
| Monocytes (Baseline) | Untreated | 9 | 12.2138 | 1.61045 | 12.4493 | 4.8314 |
| | Glatiramer | 10 | 16.0000 | 1.63379 | 16.0028 | 5.1665 |
| CD19+CD20+ (Baseline) | Untreated | 9 | 4.4065 | 0.56453 | 4.3016 | 1.6936 |
| | Glatiramer | 10 | 5.7656 | 0.87730 | 5.1053 | 2.7743 |
| CD19+CD20+CD27- Naïve (Baseline) | Untreated | 9 | 2.5904 | 0.39768 | 2.4081 | 1.1930 |
| | Glatiramer | 10 | 4.0472 | 0.77167 | 2.7997 | 2.4402 |
| CD19+CD20+CD27+ Memory (Baseline) | Untreated | 9 | 1.7378 | 0.33722 | 1.6255 | 1.0117 |
| | Glatiramer | 10 | 1.5745 | 0.18212 | 1.4892 | 0.5759 |
| CD19+CD20-CD27high Plasmablats (Baseline) | Untreated | 9 | 0.0335 | 0.00762 | 0.0292 | 0.0229 |
| | Glatiramer | 10 | 0.0596 | 0.01321 | 0.0499 | 0.0418 |
| CD19+CD20+ (Baseline, No plasmablasts) | Untreated | 9 | 4.3470 | 0.56354 | 4.2097 | 1.6906 |
| | Glatiramer | 10 | 5.6550 | 0.86954 | 4.9828 | 2.7497 |
| CD20+IgD-CD27+ Memory Switched (Baseline) | Untreated | 9 | 21.1792 | 2.71268 | 20.8879 | 8.1380 |
| | Glatiramer | 10 | 18.7718 | 2.52002 | 19.6569 | 7.9690 |
| CD20+IgD+CD27+ Memory PreSwitched (Baseline) | Untreated | 9 | 17.5969 | 2.76839 | 18.6758 | 8.3052 |
| | Glatiramer | 10 | 11.5806 | 1.53954 | 10.2769 | 4.8684 |
| CD20+IgD+CD27- Naïve (Baseline) | Untreated | 9 | 55.0889 | 4.95639 | 50.4192 | 14.8692 |
| | Glatiramer | 10 | 63.3752 | 3.88027 | 66.0922 | 12.2705 |
| CD20+IgD-CD27- Double Negative (Baseline) | Untreated | 9 | 6.1353 | 1.16227 | 4.8285 | 3.4868 |
| | Glatiramer | 10 | 6.3530 | 0.97882 | 5.8434 | 3.0953 |

| Cell Population (%) | Treatment | N | Mean | SE | Median | SD |
|---|---|---|---|---|---|---|
| Lymphocytes (12m) | Untreated | 10 | 85.8251 | 2.01614 | 87.4792 | 6.3756 |
| | Glatiramer | 10 | 84.7633 | 1.70465 | 86.6657 | 5.3906 |
| CD3+ (12m) | Untreated | 10 | 75.4878 | 3.16059 | 74.7352 | 9.9947 |
| | Glatiramer | 10 | 75.4105 | 1.50023 | 75.2325 | 4.7441 |
| CD4+ (12m) | Untreated | 10 | 47.6431 | 2.71158 | 46.2702 | 8.5748 |
| | Glatiramer | 10 | 52.2352 | 2.40317 | 52.0727 | 7.5995 |
| CD4+CD38+ (12m) | Untreated | 10 | 7.2508 | 1.17735 | 5.5658 | 3.7231 |
| | Glatiramer | 10 | 7.0554 | 1.13698 | 7.2854 | 3.5954 |
| CD4+HLADR+ (12m) | Untreated | 10 | 1.3384 | 0.12594 | 1.2536 | 0.3983 |
| | Glatiramer | 10 | 1.8958 | 0.16436 | 2.0189 | 0.5197 |
| CD4+CD38+HLADR+ (12m) | Untreated | 10 | 0.3113 | 0.05248 | 0.2596 | 0.1660 |
| | Glatiramer | 10 | 0.2984 | 0.04122 | 0.2719 | 0.1304 |
| CD4+CD25highCD127low (12m) | Untreated | 10 | 3.3967 | 0.24776 | 3.2465 | 0.7835 |
| | Glatiramer | 10 | 3.6589 | 0.30679 | 3.5444 | 0.9701 |
| Monocytes (12m) | Untreated | 10 | 14.1749 | 2.01614 | 12.5208 | 6.3756 |
| | Glatiramer | 10 | 15.2367 | 1.70465 | 13.3343 | 5.3906 |
| CD19+CD20+ (12m) | Untreated | 10 | 5.7591 | 0.91785 | 5.0998 | 2.9025 |
| | Glatiramer | 10 | 7.5630 | 0.79693 | 7.1895 | 2.5201 |
| CD19+CD20+CD27- Naïve (12m) | Untreated | 10 | 3.5575 | 0.70529 | 2.8592 | 2.2303 |
| | Glatiramer | 10 | 5.5806 | 0.76651 | 5.1331 | 2.4239 |
| CD19+CD20+CD27+ Memory (12m) | Untreated | 10 | 2.0879 | 0.38210 | 1.7902 | 1.2083 |
| | Glatiramer | 10 | 1.8316 | 0.21854 | 1.8507 | 0.6911 |
| CD19+CD20-CD27high Plasmablasts (12m) | Untreated | 10 | 0.0413 | 0.01094 | 0.0353 | 0.0346 |
| | Glatiramer | 10 | 0.0588 | 0.01559 | 0.0463 | 0.0493 |
| CD19+CD20+ Non Plasmablasts (12m) | Untreated | 10 | 5.6806 | 0.90611 | 5.0360 | 2.8654 |
| | Glatiramer | 10 | 7.4443 | 0.79652 | 6.9892 | 2.5188 |
| CD20+IgD-CD27+ Memory Switched (12m) | Untreated | 10 | 19.3871 | 2.56476 | 18.9367 | 8.1105 |
| | Glatiramer | 10 | 15.0433 | 2.01952 | 14.2399 | 6.3863 |
| CD20+IgD+CD27+ Memory PreSwitched (12m) | Untreated | 10 | 18.0298 | 2.73232 | 15.9306 | 8.6404 |
| | Glatiramer | 10 | 12.0758 | 2.08285 | 11.8274 | 6.5866 |
| CD20+IgD+CD27- Naïve (12m) | Untreated | 10 | 57.1868 | 4.79672 | 59.5811 | 15.1686 |
| | Glatiramer | 10 | 67.5421 | 3.76479 | 70.5143 | 11.9053 |
| CD20+IgD-CD27- Double Negative (12m) | Untreated | 10 | 5.3963 | 0.67378 | 5.0342 | 2.1307 |
| | Glatiramer | 10 | 5.3388 | 0.96283 | 4.3846 | 3.0447 |
| Lymphocytes (24m) | Untreated | 10 | 87.9137 | 1.11212 | 87.4664 | 3.5168 |
| | Glatiramer | 10 | 85.4291 | 1.54685 | 83.5509 | 4.8916 |
| CD3+ (24m) | Untreated | 10 | 76.7153 | 2.71842 | 76.2855 | 8.5964 |
| | Glatiramer | 10 | 71.9719 | 2.31649 | 71.4847 | 7.3254 |

| Cell Population (%) | Treatment | N | Mean | SE | Median | SD |
|---|---|---|---|---|---|---|
| CD4+ (24m) | Untreated | 10 | 48.2311 | 2.55333 | 47.7134 | 8.0743 |
| | Glatiramer | 10 | 48.9324 | 2.51619 | 49.8149 | 7.9569 |
| CD4+CD38+ (24m) | Untreated | 10 | 7.2268 | 1.25677 | 6.4953 | 3.9743 |
| | Glatiramer | 10 | 6.6929 | 1.16544 | 7.2463 | 3.6854 |
| CD4+HLADR+ (24m) | Untreated | 10 | 1.4577 | 0.18813 | 1.1937 | 0.5949 |
| | Glatiramer | 10 | 2.0291 | 0.18618 | 2.0337 | 0.5888 |
| CD4+CD38+HLADR+ (24m) | Untreated | 10 | 0.2981 | 0.04001 | 0.2810 | 0.1265 |
| | Glatiramer | 10 | 0.2601 | 0.02053 | 0.2599 | 0.0649 |
| CD4+CD25highCD127low (24m) | Untreated | 10 | 3.2104 | 0.22249 | 2.9355 | 0.7036 |
| | Glatiramer | 10 | 3.2429 | 0.36600 | 3.0308 | 1.1574 |
| Monocytes (24m) | Untreated | 10 | 12.0863 | 1.11212 | 12.5336 | 3.5168 |
| | Glatiramer | 10 | 14.5709 | 1.54685 | 16.4491 | 4.8916 |
| CD19+CD20+ (24m) | Untreated | 10 | 5.4659 | 0.70731 | 5.5267 | 2.2367 |
| | Glatiramer | 10 | 7.8512 | 0.84220 | 7.0958 | 2.6633 |
| CD19+CD20+CD27- Naïve (24m) | Untreated | 10 | 3.2494 | 0.47700 | 2.7588 | 1.5084 |
| | Glatiramer | 10 | 5.9870 | 0.79829 | 5.3351 | 2.5244 |
| CD19+CD20+CD27+ Memory (24m) | Untreated | 10 | 2.1005 | 0.38931 | 1.8149 | 1.2311 |
| | Glatiramer | 10 | 1.7009 | 0.15553 | 1.6599 | 0.4918 |
| CD19+CD20-CD27high Plasmablasts (24m) | Untreated | 10 | 0.0504 | 0.01268 | 0.0285 | 0.0401 |
| | Glatiramer | 10 | 0.0797 | 0.03909 | 0.0463 | 0.1236 |
| CD19+CD20+ No Plasmablasts (24m) | Untreated | 10 | 5.3894 | 0.71073 | 5.4833 | 2.2475 |
| | Glatiramer | 10 | 7.7199 | 0.85533 | 7.0126 | 2.7048 |
| CD20+IgD-CD27+ Memory Switched (24m) | Untreated | 10 | 19.9960 | 2.56953 | 21.0089 | 8.1256 |
| | Glatiramer | 10 | 13.3819 | 2.62634 | 12.1846 | 8.3052 |
| CD20+IgD+CD27+ Memory PreSwitched (24m) | Untreated | 10 | 19.5831 | 2.72557 | 16.3556 | 8.6190 |
| | Glatiramer | 10 | 10.4485 | 1.51220 | 9.6176 | 4.7820 |
| CD20+IgD+CD27- Naïve (24m) | Untreated | 10 | 55.4410 | 4.80968 | 55.1520 | 15.2095 |
| | Glatiramer | 10 | 71.1666 | 4.49084 | 75.0904 | 14.2013 |
| CD20+IgD-CD27- Double Negative (24m) | Untreated | 10 | 4.9366 | 0.64368 | 4.6223 | 2.0355 |
| | Glatiramer | 10 | 5.0030 | 0.79571 | 5.1136 | 2.5163 |

#### Supplement Figure 4

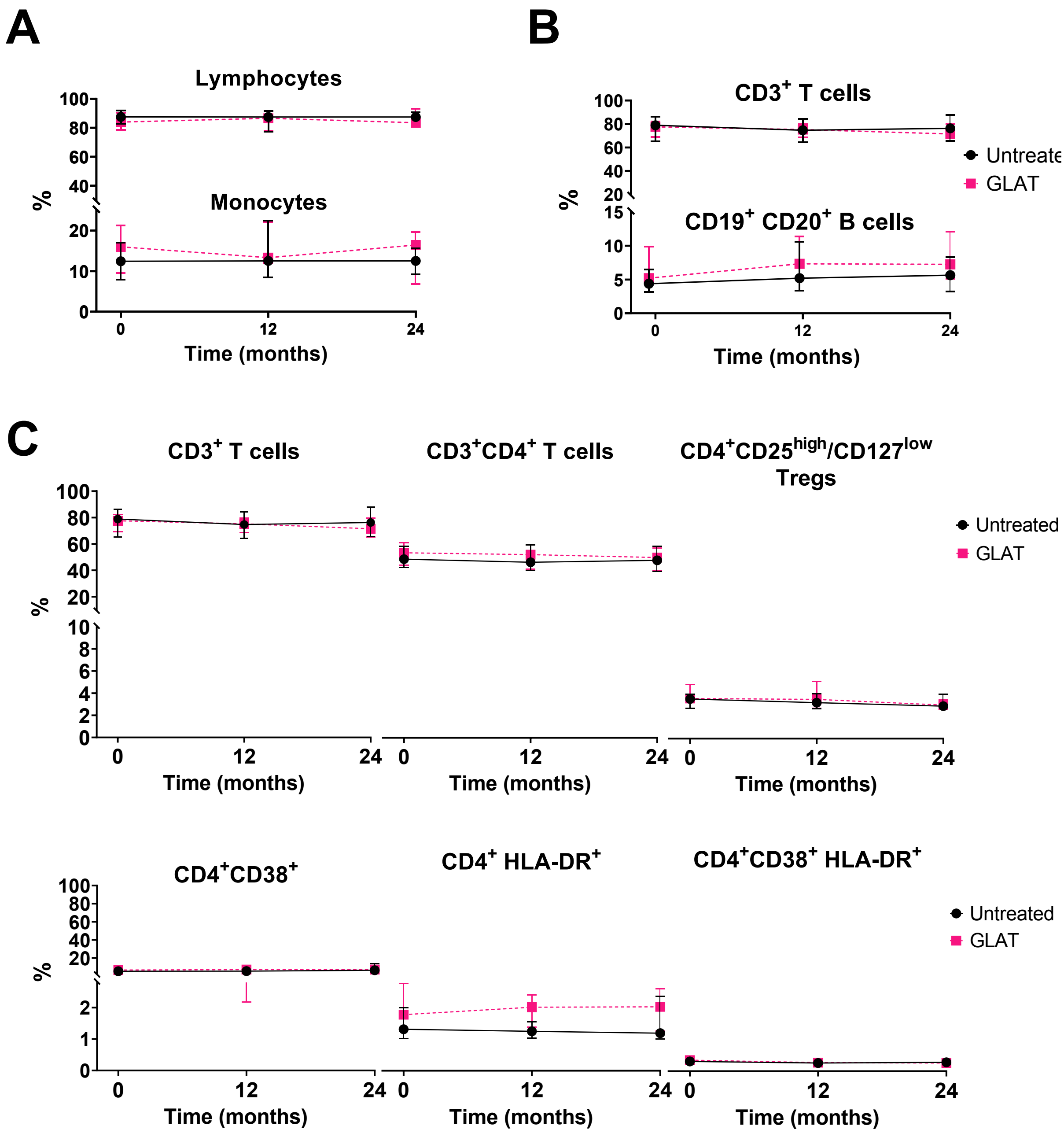


**Supplement Figure 4. Glatiramer acetate in pwCIS's peripheral lymphocytes.** A) Median percentage of lymphocytes and monocytes B) Median percentage of $CD3^+$ T cells and $CD19^+CD20^+$ B cells in the lymphocyte population. C) Median percentage of different T cell subpopulations in the lymphocyte population. Data are presented as median percentage 95% CI.

#### Supplement Figure 5

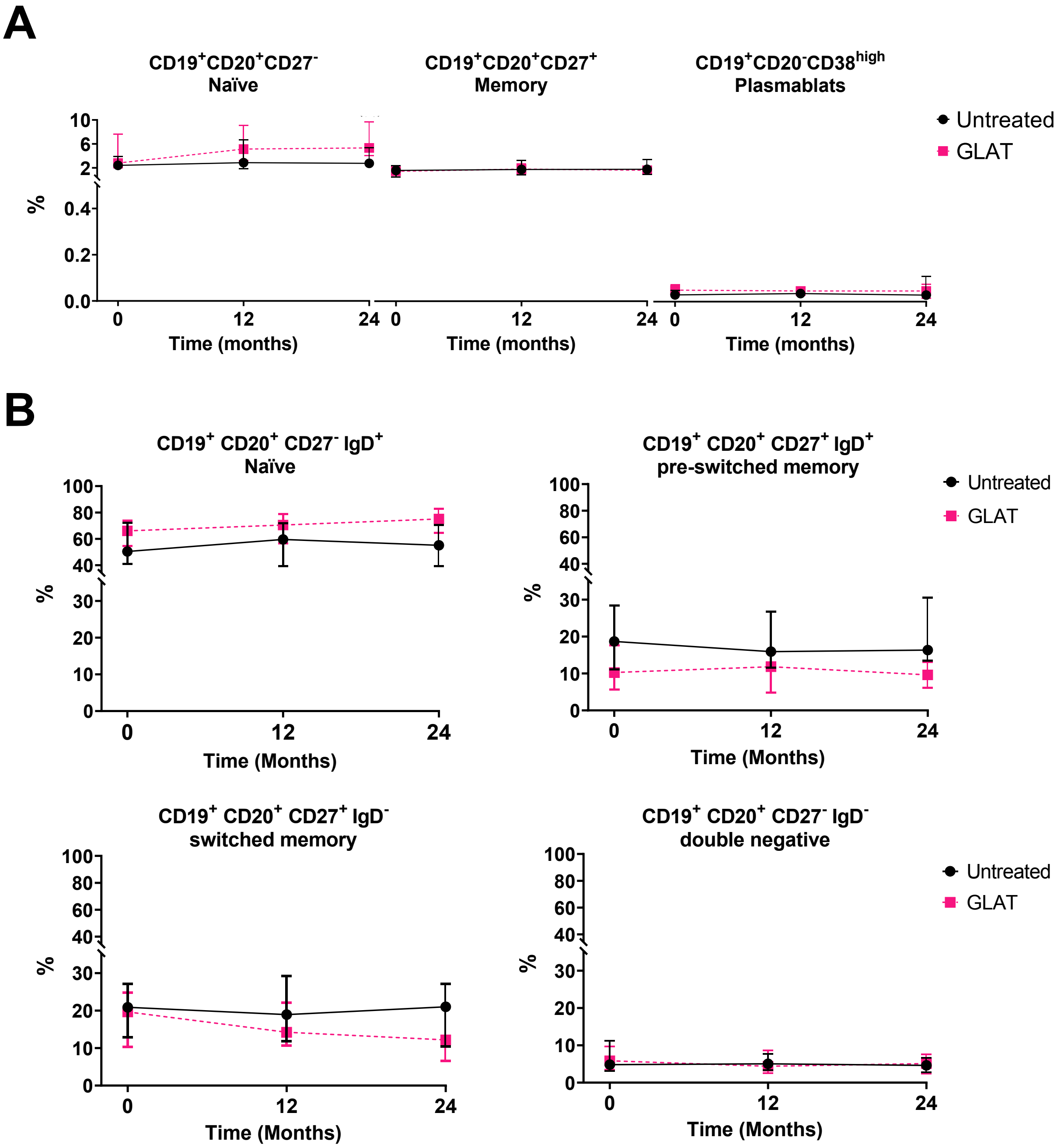


**Supplement Figure 5. Glatiramer acetate effect in pwCIS's peripheral B cells.**

A) Median percentage of different B cell subpopulations in the lymphocyte population. B) Median percentage of different B cell subpopulations into the $CD20^+$ subpopulation. Data are presented as median percentage 95% CI.
